# Snapshot PCR Surveillance for SARS-CoV-2 in Hospital Staff in England

**DOI:** 10.1101/2020.06.14.20128876

**Authors:** Colin Brown, Kathryn Clare, Meera Chand, Julie Andrews, Cressida Auckland, Sarah Beshir, Saher Choudhry, Kerrie Davies, Jane Freeman, Andrew Gallini, Rachel Moores, Trupti Patel, Gosia Poznalska, Alison Rodger, Stella Roberts, Christopher Rooney, Mark Wilcox, Simon Warren, Joanna Ellis, Robin Gopal, Jake Dunning, Maria Zambon, Susan Hopkins

**Author notes:** (co-first).

## Abstract

**Background:** Significant nosocomial transmission of SARS-CoV-2 has been demonstrated. Understanding the prevalence of SARS-CoV-2 carriage amongst HCWs at work is necessary to inform the development of HCW screening programmes to control nosocomial spread.

**Methods:** Cross-sectional ‘snapshot’ survey from April-May 2020; HCWs recruited from six UK hospitals. Participants self-completed a health questionnaire and underwent a combined viral nose and throat swab, tested by Polymerase Chain Reaction (PCR) for SARS-CoV-2 with viral culture on majority of positive samples.

**Findings:** Point prevalence of SARS-CoV-2 carriage across the sites was 2·0% (23/1152 participants), median cycle threshold value 35·70 (IQR:32·42-37·57). 17 were previously symptomatic, two currently symptomatic (isolated anosmia and sore throat); the remainder declared no prior or current symptoms. Symptoms in the past month were associated with threefold increased odds of testing positive (aOR 3·46, 95%CI 1·38-8·67; p=0·008). SARS-CoV-2 virus was isolated from only one (5%) of nineteen cultured samples. A large proportion (39%) of participants reported symptoms in the past month.

**Interpretation:** The point-prevalence is similar to previous estimates for HCWs in April 2020, though a magnitude higher than in the general population. Based upon interpretation of symptom history and testing results including viral culture, the majority of those testing positive were unlikely to be infectious at time of sampling. Development of screening programmes must balance the potential to identify additional cases based upon likely prevalence, expanding the symptoms list to encourage HCW testing, with resource implications and risks of excluding those unlikely to be infectious with positive tests.

**Funding:** Public Health England.

**Word Count:** *Research in context:* Evidence before this study
A search of PubMed was performed on 29^th^ April 2020 to identify other major works in this field, using the search terms (“novel coronavirus” OR “SARS-CoV-2” OR “COVID-19” OR “coronavirus”) AND (“workers” OR “staff”) AND (“testing” OR “screening”) from 31^st^ December 2019 onwards with no other limits. This search was updated on 10^th^ May 2020, and in addition reference lists were checked and pre-print papers were shared with us through professional networks. We found three papers commenting on prevalence of asymptomatic/pauci-symptomatic SARS-CoV-2 infection in healthcare workers, with prevalence estimates ranging from 1·1 to 8%. One of these studies explored previous symptoms in depth, though this was based upon a retrospective questionnaire and thus subject to recall bias. None of these studies explored exposures to the SARS-CoV-2 virus, commented on whether participants had been tested prior to the start of the study, or broke down results by staff role. Only one reported on estimated viral load (as inferred from cycle threshold [Ct] value), and none reported attempting viral culture. Added value of this study
This is the first published study of which we are aware that has been conducted across multiple sites in England and is therefore potentially more representative of the overall prevalence of SARS-CoV-2 infectivity amongst HCWs in the workplace. We explored symptoms in the preceding month in more depth than previous studies and in addition asked about previous test results and various exposures, also not commented on in other studies. Additionally, we attempted to isolate virus from some PCR-positive samples to look for evidence of infectious virus. Implications of all the available evidence
Authors of previous studies have proposed that screening asymptomatic HCWs for SARS-CoV-2 RNA may be beneficial, in addition to screening symptomatic HCWs. Our findings suggest that when prevalence of COVID-19 is very low, routine and repeated screening would be unlikely to have significant value, especially given the majority of participants testing positive in this study were unlikely to be infectious. However, in situations where prevalence levels are high in a particular population or setting, for example in a hospital outbreak, widening the case definition, or screening all HCWs irrespective of symptoms, may be of benefit.

## Introduction

On 31^st^ December 2019, a cluster of undiagnosed pneumonia cases was reported in Wuhan, China. The causative virus, SARS-CoV-2, was identified in January 2020, and rapidly spread through China and across the globe. By 5^th^ June 2020, there were six and a half million (6,535,354) confirmed cases of 2019 novel coronavirus disease (COVID-19). ^1^Although HCWs in the UK do not appear to be at increased risk of dying from COVID-19,^2^ health and social care workers in patient-facing or resident-facing roles appear to have higher rates of infection; between 26 April and 30 May 2020, 1·87% tested positive for COVID-19 (95% CI: 1·07%-3·02%), compared to 0·32% (95% CI: 0·26% to 0·44%) of those of working age and not in such roles.^3^ Screening of symptomatic staff in two NHS Trusts (Sheffield and Newcastle) in March 2020 demonstrated SARS-CoV-2 positivity rates of 14% and 18%.^4,5^ Significant nosocomial transmission has also been shown in a hospital in China.^6^ As local community transmission rates fall, nosocomial infection of inpatients and HCWs will likely be of increasing relative importance than infection imported from the community.^7^ In the UK, the proportion of hospitalised COVID-19 patients who developed symptoms 7 days after admission (average incubation period 5-6 days^8^) has been increasing, reaching 20% in May 2020.^9^

Estimates of the proportion of general COVID-19 cases that are asymptomatic range from 5 to 80%,^10^ with one systematic review estimating the upper bound of asymptomatic infection to be 29%.^11^ The contribution and mechanisms of asymptomatic and pre-symptomatic transmission remain unclear. Serial throat swab-sampling of Chinese COVID-19 patients suggested the infectious peak to be either prior to or at the time of symptom onset.^12^

A London hospital study found HCW prevalence to decrease from 7·1% to 1·1% over a five-week period from late March 2020; no symptoms in the week preceding or following the test were reported by around a quarter (27%) of HCWs testing positive.^13^ Screening of East of England HCWs in April 2020 found a 3·0% prevalence, with 40% of PCR-positive HCWs reporting previous symptoms (at least 7 days prior to testing.)^14^ Viral loads were significantly higher in symptomatic HCWs excluded from, compared to those remaining at, work. Screening in a Midlands hospital, also in April, reported a prevalence of 2·4%; a quarter had previous symptoms.^15^

To inform the design of HCW screening programmes, we aimed to ascertain a snapshot of the proportion of working hospital staff in whom SARS-CoV-2 could be detected, how this relates to exposure histories and reported symptoms prior to sampling, and if virus could be isolated from PCR-positive samples, indicate ng increased onward risk of infection to others.

## Methods

### Study design, setting and participants

For this prospective, cross-sectional, multi-centre study performed as a public health investigation as part of Public Health England’s national incident response to COVID-19, staff from six hospitals were invited to participate from 24^th^ April 2020 to 7^th^ May 2020. Ethics approval (NR0202) was obtained from the PHE Research Support and Governance Office prior to commencement and for all protocol changes.

The hospitals selected were a convenience sample located in London and north and south England, encompassing five NHS Trusts and one an independent hospital with a charitable hospice.

Swabbing occurred at each site over one to two days. All staff cadres present on the day of testing were eligible to participate. Local site investigators visited different hospital areas to recruit HCWs from a range of job roles.

Staff who volunteered to participate provided informed consent and were allocated a unique identifier. For the first four sites, the protocol prescribed full anonymisation; participants were consented not to be given their results and each participant’s unique identifier was recorded on the specimen and questionnaire, but not linked to any personally identifiable information and was not recorded on the consent form, nor revealed to the participant. Full-anonymisation was based upon the lack of consensus regarding the significance of the detection of viral RNA in a HCW not meeting the national case definition, how such cases and their contacts should be managed, and to allow staff participation without concern of reporting symptoms.

For the latter two sites, participants provided consent to be given positive and negative results by their respective employers; this approved protocol amendment change reflected an emerging national consensus around recommendations for testing asymptomatic NHS hospital staff. Staff with positive PCR results were advised according to local policies. The unique participant number was recorded on the consent form, kept by the local site, to allow positive results to be matched and reported to participants. At two sites, asymptomatic participants with positive PCR results were also followed up to monitor for development of symptoms.

### Procedures

Each participant was asked to complete a one-page questionnaire requesting information about demographics; job role and area of work; whether this involves aerosol generating procedures; personal and household symptom history in the past month, current symptoms, seasonal allergy symptoms; previous testing for SARS-CoV-2 and exposures to confirmed or suspected COVID-19 cases without PPE either at work or in the community (see Appendix 1). Responses were subsequently entered into a database by Public Health England (PHE) staff.

A combined viral throat and nose swab was taken from each participant by experienced staff and placed in viral transport medium, respecting local Trust infection prevention and control and PPE requirements. These were transferred to the laboratory on the same day for detection of SARS-CoV-2 RNA by real-time reverse transcription Polymerase Chain Reaction (PCR) at the PHE national reference laboratory (five hospitals) or one hospital laboratory. The PHE laboratory used an Applied Biosystems 7500 FAST system targeting a conserved region of the SARS-CoV-2 open reading frame (ORF1ab) gene. The hospital laboratory used a CE-IVD kit GeneFinder™ COVID-19 Plus RealAmp Kit by OSANGHealtcare, on the ELITe InGenius® platform, targeting 3 SARS-CoV-2 genes (RdRp, E, and N). Both PCRs had internal controls. Viral culture of PHE laboratory positives was attempted in Vero E6 cells with virus detection confirmed by cytopathic effect up to 14 days post-inoculation.

### Study size

A sample size calculation estimated 98 staff would require testing to detect a 10% prevalence (based upon unpublished data in early to mid-April 2020 shared through professional networks) with a margin of error 5% (with 90% power).

### Statistical analysis

Data was cleaned, with rules and changes agreed between two investigators. Stata (version 15, StataCorp, Texas) was used to describe the data and a random effects regression analysis was performed to examine the relationship between positive results and demographics and *a priori* exposures that were thought predictive of infection. Variables with *p* ≤ 0·1 in univariate analyses were included in random effects multivariate model.

## Results

### Demographics

Across the 6 sites, a total of 1,152 staff were recruited. Participants included clinical and support staff working in various locations across the sites, representing a diverse range of job roles. Demographics are presented in Table 1. Almost all (93%) stated that they worked in a patient-facing environment; 20% worked directly on COVID-19 wards; and 38% said their work involved performing aerosol generating procedures (AGPs). The top five groups of staff included were nurses (30%), doctors (20%), occupational therapists and physiotherapists (7%), other allied clinical staff (6%) and cleaners (6%). Seventy percent of participants were female. The median age was 39 years (range 19-68 years old) and one-third (32%) reported that they were from black, Asian and minority ethnic (BAME) backgrounds.

**Table 1.**
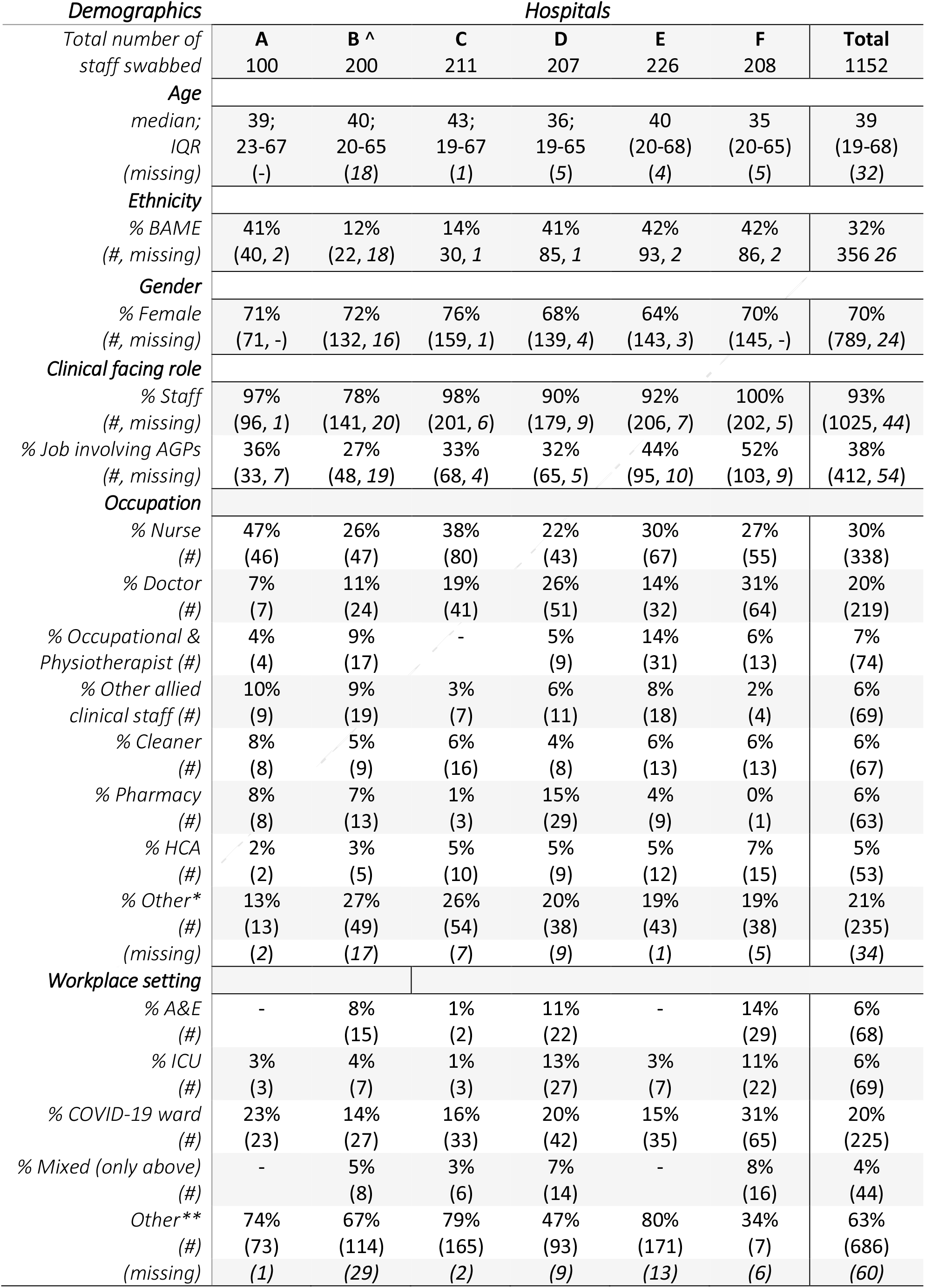
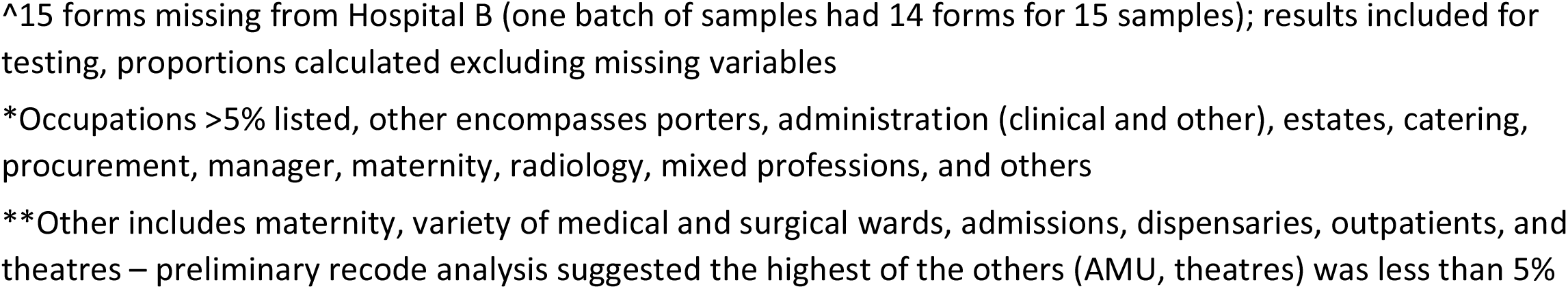
Staff demographics.

| **Demographics** | **Hospitals** |  |  |  |  |  |  |
| --- | --- | --- | --- | --- | --- | --- | --- |
| *Total number of staff swabbed* | **A** | **B ^** | **C** | **D** | **E** | **F** | **Total** |
|  | 100 | 200 | 211 | 207 | 226 | 208 | 1152 |
| **Age** |  |  |  |  |  |  |  |
| *median;* | 39; | 40; | 43; | 36; | 40 | 35 | 39 |
| *IQR* | 23-67 | 20-65 | 19-67 | 19-65 | (20-68) | (20-65) | (19-68) |
| *(missing)* | (-) | (18) | (1) | (5) | (4) | (5) | (32) |
| **Ethnicity** |  |  |  |  |  |  |  |
| *% BAME* | 41% | 12% | 14% | 41% | 42% | 42% | 32% |
| *(#, missing)* | (40, 2) | (22, 18) | 30, 1 | 85, 1 | 93, 2 | 86, 2 | 356 26 |
| **Gender** |  |  |  |  |  |  |  |
| *% Female* | 71% | 72% | 76% | 68% | 64% | 70% | 70% |
| *(#, missing)* | (71, -) | (132, 16) | (159, 1) | (139, 4) | (143, 3) | (145, -) | (789, 24) |
| **Clinical facing role** |  |  |  |  |  |  |  |
| *% Staff* | 97% | 78% | 98% | 90% | 92% | 100% | 93% |
| *(#, missing)* | (96, 1) | (141, 20) | (201, 6) | (179, 9) | (206, 7) | (202, 5) | (1025, 44) |
| *% Job involving AGPs* | 36% | 27% | 33% | 32% | 44% | 52% | 38% |
| *(#, missing)* | (33, 7) | (48, 19) | (68, 4) | (65, 5) | (95, 10) | (103, 9) | (412, 54) |
| **Occupation** |  |  |  |  |  |  |  |
| *% Nurse* | 47% | 26% | 38% | 22% | 30% | 27% | 30% |
| *(#)* | (46) | (47) | (80) | (43) | (67) | (55) | (338) |
| *% Doctor* | 7% | 11% | 19% | 26% | 14% | 31% | 20% |
| *(#)* | (7) | (24) | (41) | (51) | (32) | (64) | (219) |
| *% Occupational & Physiotherapist* | 4% | 9% | - | 5% | 14% | 6% | 7% |
| *(#)* | (4) | (17) |  | (9) | (31) | (13) | (74) |
| *% Other allied clinical staff* | 10% | 9% | 3% | 6% | 8% | 2% | 6% |
| *(#)* | (9) | (19) | (7) | (11) | (18) | (4) | (69) |
| *% Cleaner* | 8% | 5% | 6% | 4% | 6% | 6% | 6% |
| *(#)* | (8) | (9) | (16) | (8) | (13) | (13) | (67) |
| *% Pharmacy* | 8% | 7% | 1% | 15% | 4% | 0% | 6% |
| *(#)* | (8) | (13) | (3) | (29) | (9) | (1) | (63) |
| *% HCA* | 2% | 3% | 5% | 5% | 5% | 7% | 5% |
| *(#)* | (2) | (5) | (10) | (9) | (12) | (15) | (53) |
| *% Other\** | 13% | 27% | 26% | 20% | 19% | 19% | 21% |
| *(#)* | (13) | (49) | (54) | (38) | (43) | (38) | (235) |
| *(missing)* | (2) | (17) | (7) | (9) | (1) | (5) | (34) |
| **Workplace setting** |  |  |  |  |  |  |  |
| *% A&E* | - | 8% | 1% | 11% | - | 14% | 6% |
| *(#)* |  | (15) | (2) | (22) |  | (29) | (68) |
| *% ICU* | 3% | 4% | 1% | 13% | 3% | 11% | 6% |
| *(#)* | (3) | (7) | (3) | (27) | (7) | (22) | (69) |
| *% COVID-19 ward* | 23% | 14% | 16% | 20% | 15% | 31% | 20% |
| *(#)* | (23) | (27) | (33) | (42) | (35) | (65) | (225) |
| *% Mixed (only above)* | - | 5% | 3% | 7% | - | 8% | 4% |
| *(#)* |  | (8) | (6) | (14) |  | (16) | (44) |
| *Other\*\** | 74% | 67% | 79% | 47% | 80% | 34% | 63% |
| *(#)* | (73) | (114) | (165) | (93) | (171) | (7) | (686) |
| *(missing)* | (1) | (29) | (2) | (9) | (13) | (6) | (60) |

^15 forms missing from Hospital B (one batch of samples had 14 forms for 15 samples); results included for testing, proportions calculated excluding missing variables
\*Occupations >5% listed, other encompasses porters, administration (clinical and other), estates, catering, procurement, manager, maternity, radiology, mixed professions, and others
\*\*Other includes maternity, variety of medical and surgical wards, admissions, dispensaries, outpatients, and theatres – preliminary recode analysis suggested the highest of the others (AMU, theatres) was less than 5%

### Symptoms, prior testing and exposures

438 (39%) of all participants who answered the questions about symptoms (1125), had experienced at least one respiratory, gastrointestinal or influenza-like symptom in the previous month (see Figure 1). Of those, half (50%) reported symptoms in keeping with the national COVID-19 case definition (Table 2 details staff illness, symptoms, and exposures). Among the 426 with a test result available, headache, cough and sore throat were common across those who tested positive and negative (53% vs 52%; 53% vs 42%; and 53% vs 42%). Myalgia, anosmia, change in taste and fever were markedly more common in HCWs who tested positive (60% vs 29%, Fisher’s Exact *p*=0·019; 53% vs 19%, *p*=0·004; 47% vs· 17%, *p*=0·008; 47% vs 24%, *p*=0·056). Runny nose was more common in those who tested negative (20% versus 58%, *p*=0·185).

**Figure 1.**
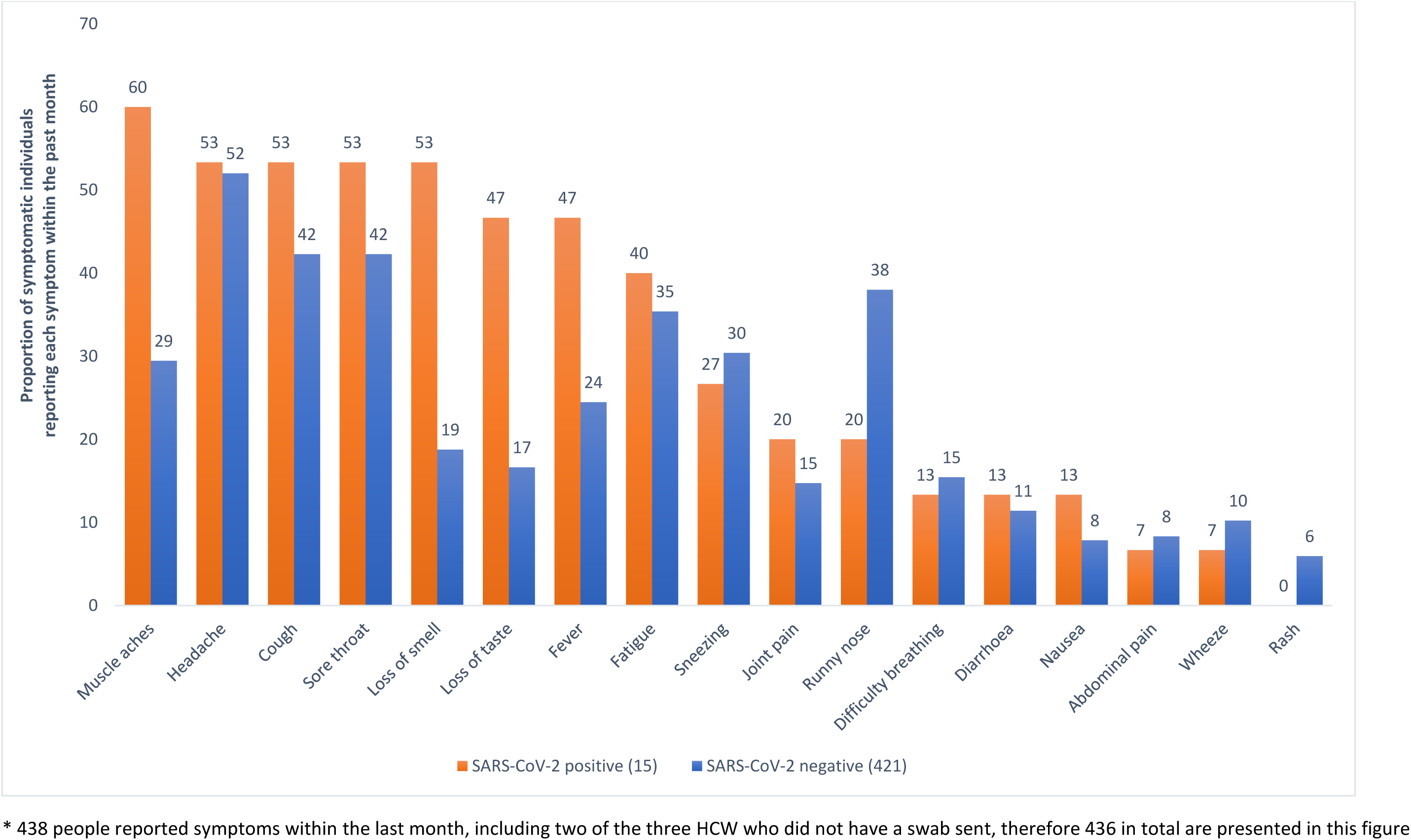
Proportion of individuals reporting symptoms in the last month by SARS-CoV-2 PCR status and symptom.

**Table 2.** Staff illness, symptoms, and exposures.

| *Illness* | *Hospitals* |  |  |  |  |  | **Total** |
| --- | --- | --- | --- | --- | --- | --- | --- |
|  | **A** | **B <sup>^</sup>** | **C** | **D** | **E** | **F** |  |
| *Total number of staff swabbed* | 100 | 200 | 211 | 207 | 226 | 208 | 1152 |
| **Symptoms** |  |  |  |  |  |  |  |
| *% Last month* | 38% | 42% | 35% | 49% | 28% | 43% | 39% |
| *(number, missing <sup>Ω</sup>)* | (38, 1) | (77, 15) | (73, 3) | (101, 1) | (63, 1) | (86, 6) | (438, 27) |
| *% Symptomatic with fever and/or cough (#)* | 58% | 41% | 36% | 55% | 65% | 51% | 50% |
| *(#)* | (21) | (32) | (26) | (56) | (41) | (44) | (220) |
| *% Current symptoms\* (#, missing)* | 31% | 26% | 28% | 22% | 22% | 24% | 25% |
| *(#, missing)* | (11, 2) | (19, 5) | (19, 5) | (22, 1) | (13, 3) | (19, 7) | (103, 23) |
| *% Current with fever and/or cough\*\* (#)* | 73% | 32% | 21% | 45% | 62% | 63% | 47% |
| *(#)* | (8) | (6) | (4) | (10) | (8) | (12) | (55) |
| **SARS-CoV-2 testing** |  |  |  |  |  |  |  |
| *% Previously tested (#, missing <sup>Ω</sup>) <sup>α</sup>* | 6% | 6% | 12% | 12% | 8% | 11% | 10% |
| *(#, missing <sup>Ω</sup>) <sup>α</sup>* | (5, 21) | (8, 89) | (15, 88) | (18, 63) | (10, 103) | (17, 54) | (71, 420) |
| *% Tested positive <sup>β</sup> (#, missing)* | 20% | 25% | 21% | 22% | 40% | 31% | 26% |
| *(#, missing)* | (1, -) | (2, -) | (3, 1) | (4, -) | (4, -) | (5, 1) | (19, 2) |
| **Household illness** |  |  |  |  |  |  |  |
| *% Respiratory symptoms last month (#, missing)* | 12% | 9% | 8% | 14% | 15% | 13% | 12% |
| *(#, missing)* | (12, 1) | (17, 17) | (16, 2) | (29, 4) | (34, 2) | (12, 1) | (134, 29) |
| *% Previously tested <sup>α</sup> (#, missing)* | 4% | 11% | 20% | 9% | 11% | 11% | 12% |
| *(#, missing)* | (4, 57) | (8, 142) | (11, 156) | (6, 139) | (9, 142) | (14, 131) | (47, 769) |
| *% Tested positive <sup>β</sup> (#, missing)* | 25% | 0% | 20% | 33% | 38% | 56% | 29% |
| *(#, missing)* | (1, -) | (0, -) | (2, 1) | (2, 1) | (3, 1) | (5, 5) | (13, 8) |
| **Seasonal symptoms** |  |  |  |  |  |  |  |
| *% Usual hayfever (#, missing)* | 43% | 41% | 40% | 40% | 36% | 39% | 39% |
| *(#, missing)* | (42, 3) | (73, 37) | (83, 39) | (80, 39) | (78, 35) | (78, 38) | (434, 52) |
| **Exposure without PPE** |  |  |  |  |  |  |  |
| *% No exposure (#)* | 49% | 75% | 75% | 55% | 57% | 56% | 62% |
| *(#)* | (47) | (133) | (158) | (112) | (108) | (112) | (670) |
| *% Community exposure (#)* | 13% | 2% | 2% | 4% | 6% | 2% | 4% |
| *(#)* | (12) | (3) | (5) | (9) | (11) | (5) | (45) |
| *% Workplace exposure (#)* | 39% | 23% | 23% | 40% | 37% | 42% | 34% |
| *(#)* | (37) | (41) | (48) | (82) | (71) | (84) | (363) |
| *(missing)* | (4) | (23) | (-) | (4) | (36) | (7) | (74) |

<sup>^</sup>15 forms missing from Hospital B (one batch of samples had 14 forms for 15 samples); results included for testing, proportions calculated excluding missing variables
\*Percentage of those who answered if they had symptoms within last month
\*\*Fever and cough initially reported, for those who reported current symptoms unclear if these are present
<sup>α</sup> Percentage of those who answered, likely influenced by presence of symptoms (but not completely)
<sup>β</sup> Percentage of those who have reported a previous test
<sup>Ω</sup> Or unclear e.g. ticked no symptoms within last month and then ticked individual symptoms

**Table 3.**
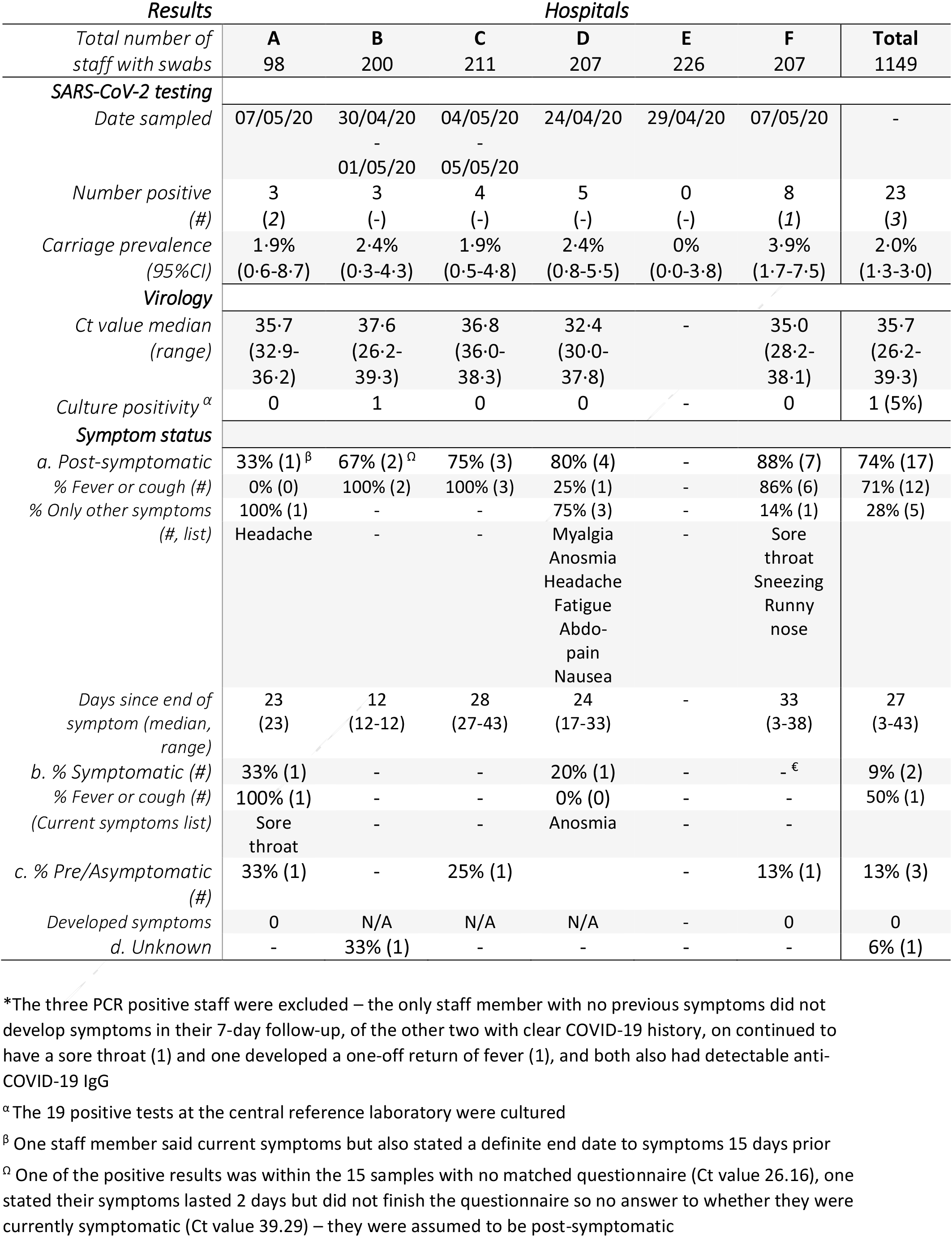
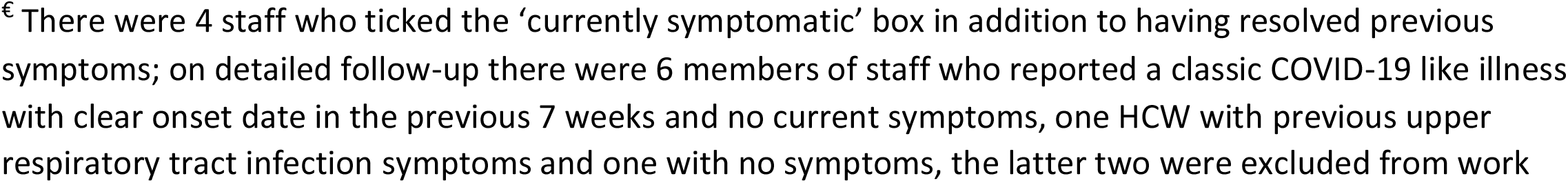
Laboratory results and follow-up.

| *Results* |  | *Hospitals* |  |  |  |  |  |
| --- | --- | --- | --- | --- | --- | --- | --- |
| *Total number of staff with swabs* | **A** | **B** | **C** | **D** | **E** | **F** | **Total** |
|  | 98 | 200 | 211 | 207 | 226 | 207 | 1149 |
| **SARS-CoV-2 testing** |  |  |  |  |  |  |  |
| *Date sampled* | 07/05/20 | 30/04/20<br>01/05/20 | 04/05/20<br>05/05/20 | 24/04/20 | 29/04/20 | 07/05/20 | - |
| *Number positive (#)* | 3<br>(2) | 3<br>(-) | 4<br>(-) | 5<br>(-) | 0<br>(-) | 8<br>(1) | 23<br>(3) |
| *Carriage prevalence (95%CI)* | 1.9%<br>(0.6-8.7) | 2.4%<br>(0.3-4.3) | 1.9%<br>(0.5-4.8) | 2.4%<br>(0.8-5.5) | 0%<br>(0.0-3.8) | 3.9%<br>(1.7-7.5) | 2.0%<br>(1.3-3.0) |
| **Virology** |  |  |  |  |  |  |  |
| *Ct value median (range)* | 35.7<br>(32.9-36.2) | 37.6<br>(26.2-39.3) | 36.8<br>(36.0-38.3) | 32.4<br>(30.0-37.8) | - | 35.0<br>(28.2-38.1) | 35.7<br>(26.2-39.3) |
| *Culture positivity<sup>α</sup>* | 0 | 1 | 0 | 0 | - | 0 | 1 (5%) |
| **Symptom status** |  |  |  |  |  |  |  |
| *a. Post-symptomatic* | 33% (1)<sup>β</sup> | 67% (2)<sup>Ω</sup> | 75% (3) | 80% (4) | - | 88% (7) | 74% (17) |
| *% Fever or cough (#)* | 0% (0) | 100% (2) | 100% (3) | 25% (1) | - | 86% (6) | 71% (12) |
| *% Only other symptoms (#, list)* | 100% (1)<br>Headache | - | - | 75% (3)<br>Myalgia<br>Anosmia<br>Headache<br>Fatigue<br>Abdo-pain<br>Nausea | - | 14% (1)<br>Sore throat<br>Sneezing<br>Runny nose | 28% (5) |
| *Days since end of symptom (median, range)* | 23<br>(23) | 12<br>(12-12) | 28<br>(27-43) | 24<br>(17-33) | - | 33<br>(3-38) | 27<br>(3-43) |
| *b. % Symptomatic (#)* | 33% (1) | - | - | 20% (1) | - | -<sup>€</sup> | 9% (2) |
| *% Fever or cough (#)* | 100% (1) | - | - | 0% (0) | - | - | 50% (1) |
| *(Current symptoms list)* | Sore throat | - | - | Anosmia | - | - |  |
| *c. % Pre/Asymptomatic (#)* | 33% (1) | - | 25% (1) |  | - | 13% (1) | 13% (3) |
| *Developed symptoms* | 0 | N/A | N/A | N/A | - | 0 | 0 |
| *d. Unknown* | - | 33% (1) | - | - | - | - | 6% (1) |

\*The three PCR positive staff were excluded – the only staff member with no previous symptoms did not develop symptoms in their 7-day follow-up, of the other two with clear COVID-19 history, one continued to have a sore throat (1) and one developed a one-off return of fever (1), and both also had detectable anti-COVID-19 IgG
<sup>α</sup> The 19 positive tests at the central reference laboratory were cultured
<sup>β</sup> One staff member said current symptoms but also stated a definite end date to symptoms 15 days prior
<sup>Ω</sup> One of the positive results was within the 15 samples with no matched questionnaire (Ct value 26.16), one stated their symptoms lasted 2 days but did not finish the questionnaire so no answer to whether they were currently symptomatic (Ct value 39.29) – they were assumed to be post-symptomatic
€ There were 4 staff who ticked the 'currently symptomatic' box in addition to having resolved previous symptoms; on detailed follow-up there were 6 members of staff who reported a classic COVID-19 like illness with clear onset date in the previous 7 weeks and no current symptoms, one HCW with previous upper respiratory tract infection symptoms and one with no symptoms, the latter two were excluded from work

One in four of all participants (25%) reporting symptoms in the past month still had symptoms on the day of sampling. We did not ask explicitly what these symptoms were, however 47% of those with current symptoms reported that they had experienced at least one of cough or fever in the past month. One in ten (12%) reported a household member with a respiratory illness in the past month.

Nearly four in ten (39%) reported seasonal respiratory allergies; there was an association between those reporting symptoms in the past month and presence of seasonal allergies (44% [188] versus 36% [234], χ2 *p*=0·006).

10% of participants who answered had been tested for SARS-CoV-2 previously, one quarter (26%) of whom had tested positive. This was similar for testing within their household (12% of those who answered had been tested, with 29% positivity).

One third of all participants (34%) reported an exposure to COVID-19 without appropriate PPE in the hospital, and this was consistent across hospital sites (range 23-42%). 4% had experienced an exposure in the community, and around one in eight (13%) of all participants had a household member who had been unwell with respiratory symptoms in the past month.

### Testing

Twenty-three out of 1152 staff (2·0%) tested positive for SARS-CoV-2. For three enrolled participants no sample was received in the laboratory. Test positivity ranged from 0% to 3·9% across the sites.

Ct values ranged from 26·2 to 39·3, with a median of 35·7 (IQR:32·42-37·57, lower Ct values indicate larger amounts of viral RNA). Viral culture was completed for all nineteen samples that were PCR positive in the central PHE laboratory. SARS-CoV-2 virus was isolated from only one sample (with a Ct value of 26·2); this was one of 15 samples unable to be matched to a questionnaire from one site.

Amongst the PCR positive participants, there was one missing and one incomplete questionnaire. Of those testing positive and for whom data were available, seventeen (74% of all positive participants) had experienced previous symptoms, with a median time from end of symptoms to sampling of 27 days (range 3-43). Thirteen (68% of all 19 participants reporting symptoms) had experienced symptoms compatible with the national case definition at the time for COVID-19 i.e. cough or fever. Five (22%) disclosed that they had had a previous test, and of these four (17%) had tested positive.

In regression analysis, staff reporting previous symptoms in the last month had markedly increased odds of testing positive (aOR 3·31, 95%CI 1·33-8·26; *p*=0·008), adjusted for age and gender (see Table 4).

**Table 4.** Random effects regression analysis.

|  | Proportion positive<br>(#/23,<br>missing<sup>α</sup>) | Proportion* negative<br>(#/1126,<br>missing<sup>α</sup>) | Univariate analysis |  | Multivariate analysis<br>(with age and gender) |  |
| --- | --- | --- | --- | --- | --- | --- |
|  |  |  | Odds Ratio (95% CI)<sup>a</sup> | p value<sup>a</sup> | Odds Ratio (95% CI)<sup>a</sup> | p value<sup>a</sup> |
| Age (mean) | 41.2<br>(1) | 39.9<br>(32) | 1.01 (0.98-1.05) per increasing year | 0.456 | 1.02 (0.98-1.06) | 0.328 |
| Gender (female) | 36%<br>(8, 1) | 30%<br>(331, 23) | 0.73 (0.30-1.78)<br>for females | 0.494 | 0.57 (0.28-1.65) | 0.389 |
| Ethnicity (BAME) | 32%<br>(7, 1) | 32%<br>(348, 25) | 0.96 (0.38-2.42)<br>for non-BAME staff | 0.921 |  |  |
| Clinical role | 90%<br>(18, 3) | 93%<br>(81, 41) | 1.99 (0.41-9.72)<br>for non-clinical jobs | 0.396 |  |  |
| Perform AGPs | 42%<br>(8, 4) | 37%<br>(403, 49) | 0.86 (0.34-2.18)<br>for not performing AGPs | 0.750 |  |  |
| Symptoms in last month | 68%<br>(15, 1) | 38%<br>(421, 16) | 3.31 (1.33-8.26)<br>for those with symptoms last month | 0.010 | 3.46 (1.38-8.67) | 0.008 |
| Case definition symptoms | 66%<br>(10, 0) | 50%<br>(170, 0) | 2.01 (0.67-5.98) if fever and/or cough | 0.210 |  |  |
| Seasonal allergies | 29%<br>(6, 2) | 40%<br>(426, 50) | 1.67 (0.64-4.34)<br>for those who do not have allergies | 0.296 |  |  |
| Current symptoms<sup>α</sup> | 21%<br>(3, 1) | 25%<br>(100, 21) | 0.82 (0.22-2.99)<br>for those with current symptoms | 0.762 |  |  |
| Household symptoms | 19%<br>(4, 2) | 14%<br>(146, 58) | 1.26 (0.47-3.37)<br>for those with symptomatic households | 0.639 |  |  |
| Work exposure (with no PPE)<sup>β</sup> | 35%<br>(7, 3) | 34%<br>(356, 71) | 1.08 (0.42-2.76)<br>for healthcare exposures | 0.881 |  |  |

\* Unless stated otherwise
<sup>α</sup> Percentage of those who answered if they had symptoms within past month
<sup>a</sup> $p$ values calculated using Wald tests, odds ratios calculated by logistic regression employing random effects
<sup>β</sup> No positives with community exposure
<sup>Ω</sup> Or unclear e.g. ticked no symptoms within last month and then ticked individual symptoms

Only two participants who tested positive were symptomatic on the day of testing; one had experienced isolated anosmia for seven days and had previously tested SARS-CoV-2 negative, one had experienced multiple symptoms (including cough and anosmia) and remained off work for 14 days, and had isolated sore throat remaining on testing 28 days after symptom onset. Four staff who had no clinical symptoms either prior to or at the time of testing were followed-up over the next week; none developed symptoms.

## Discussion

### Interpretation of results

We found relatively low rates of SARS-CoV-2 carriage in HCWs at work; 2·0% across all sites (95% CI 1·3-3·0%), with prevalence ranging from 0% to 3·9%. This is in keeping with other UK hospital estimates from a similar time period (one study identified by our literature review found prevalence to fall from 4·9% to 1·1% over April,^13^ and the other studies falling within this range).^14,15^ This is eight-fold higher than the general population (0·24%) during a similar time-period,^16^ and likely higher due to additional symptomatic HCWs self-isolating, though this snap-shot coincided with steep reductions in COVID-19 incidence.^17^ The observed differences across sites likely arise from differing community prevalence, hospital transmission rates, infection prevention and control measures, and small individual numbers. The one hospital with no positive HCWs was a ‘clean’ specialist referral centre with no emergency COVID-19 admissions.

PCR testing detects viral RNA and positivity does not necessarily indicate infectious virus. Those with mild or asymptomatic infection may be less infectious than those with respiratory symptoms, given the commonly accepted natural transmission routes for COVID-19 (respiratory droplet and direct and indirect contact routes).^18^ Those who remain SARS-CoV-2 positive on testing after resolution of symptoms i.e. people on the tail end of PCR positivity, will not necessarily be infectious. Viral RNA shedding has been reported up to 49-60 days after onset of illness,^19,20^ whereas the longest duration of detection of culturable virus is 8-9 days.^21,22^

The Ct values of 23 staff who tested SARS-CoV-2 positive in our study suggest the majority had a low viral load and were unlikely to be infectious at time of testing (see Supplement S1). SARS-CoV-2 virus was isolated only from the sample with the lowest Ct value (26·2); this mirrors a Canadian study where SARS-CoV-2 was only cultured from 90 samples where the Ct value was less than 24.^23^

Modelling from April 2020 predicted that if both asymptomatic and symptomatic HCWs were screened and isolated, transmission could be further reduced by one third (depending on timeliness of results).^18^ Only four HCWs in this cohort had detectable viral RNA without reporting either current or previous symptoms (0·3%), implying that rates of asymptomatic or pre-symptomatic COVID-19 infections were low. Therefore, the impact of mass HCW screening would be reduced, and we suggest that current prevalence estimates from surveillance data should inform when to institute HCW screening programmes.

Five (22%) of 23 positive HCWs experienced symptoms not in the national case definition; the national case definition was amended on May 18^th^ 2020 to include a loss or change in sense of smell or taste, however three cases (13%) complained only of myalgia, fatigue, headache and gastro-intestinal symptoms, assuming they accurately recalled symptoms. During times of higher prevalence, such as in a hospital outbreak, the number of such cases would be higher and using a broader case HCW definition would capture more cases; this would need to be weighed against resource implications and risks associated with false positives in lower prevalence settings.^24^ As we enter summer, the prevalence of other non-COVID respiratory illnesses is lower, and the predictive value of milder symptoms may increase, though this could be offset by hayfever prevalence. We found a strong association with myalgia and a positive test – this was among the most reported symptoms (57%) in a French case series of nearly 1500 patients after fever and cough,^25^ though was much less common in a large UK study of over 20,000 inpatients (∼20%).^26^

One third of participants reported an exposure without appropriate PPE in the hospital setting, though we did not capture the details of these such as PPE breaches or inadvertent exposures to HCWs or patients. It is not possible to infer whether acquisition was community or nosocomial.

### Strengths and limitations

This study is the first which estimates point prevalence of SARS-CoV-2 carriage in HCWs at work in multiple job roles across a number of hospitals in different UK regions. Our study also includes a larger number of participants than previous reported studies. Prevalence among HCWs will be dynamic, and likely to change as the infection rate across the whole population falls.^13^ This snapshot study is unable to capture such trends.

Selection bias could be present in either direction, particularly for the latter two sites: staff may have been more inclined to volunteer if they were concerned about COVID-19 infection, or less likely if they were anxious of work exclusion for themselves or their household. Staff with symptoms or exposures may have been less inclined to report these honestly (information bias), though reassurance about confidentiality will have at least in part mitigated this. The potential for symptom and exposure recall bias about was present throughout, and questionnaires were single data entered.

Due to full anonymisation we were unable to follow-up one participant who was asymptomatic at the time of testing to see if they developed symptoms, and the one culture positive participant was unable to be matched to a questionnaire, so it is not possible to estimate the true asymptomatic infection prevalence. Of the 17 post-symptomatic HCWs, four had a previous positive test result, and a further six were followed-up at the two latter hospital sites; the remaining seven were assumed to be post-symptomatic (all but one had fever, cough or anosmia).

## Conclusion

Although the point-prevalence of SARS-CoV-2 RNA detection is higher for our HCW population than for the general population, the majority of HCWs identified were unlikely to be infectious at the time of testing, based on Ct values, viral culture results and symptom history. Testing for SARS-CoV-2 in HCWs who meet the current national case definition of cough, fever or change in sense of taste or smell may not capture all positive cases. Screening HCWs based on a broader case definition, or whilst asymptomatic in certain situations, may be more beneficial when community prevalence is rising or high; however screening should balance the benefit of identifying additional cases against the resource implications and the risk of excluding staff who are not infectious.

## Author contributions

CB, KC, MC, JD, MZ & SH designed the study; JA, CA, SB, SC, KD, JF, AG, RM, TP, GP, AR, SR, CR, MW, SW & JD arranged the administration of the questionnaires and swabbing of the HCW; CA, JE, RG & MZ arranged for laboratory testing of the samples; CB, KC, MC, JD & SH wrote the initial manuscript draft; JA, AR, VA, KD, RM, TP, AR & MW gave initial substantive comments; all authors reviewed and agreed the final text.

## Data Availability

Data available on request

## Acknowledgements

Public Health England (Mary Ramsay, Vanessa Saliba, Julie Robotham, Rifat Sofoo, Maria Kephalas, Mehdi Minaji, Joanna Connelly, Nandini Shetty, Steve Harbour, Lynne Foster, Gwyn Morris), Whittington Hospital (Helena Rochford, Joseph Grant, Matt Kinsella, Becky Lai, Neil Jones, Naina McCann), Leeds Teaching Hospital Trust (Phillip Wood), Royal Marsden Hospital (Pat Cattini), Royal National Orthopaedic Hospital (Iva Hauptmannova, Deidre Brooking, Andrew Symonds, Esther Hanison), Royal Devon & Exeter NHS Foundation Trust (Helen Quinn, Stephanie Estcourt, RDE Research & Development Team), Hospital of St John & St Elizabeth (Jozanne Bates, Giovanni D Stefano), Royal Free Hospital (Stephen Mepham, Clare Warrell, Aoife Malloy, Tehmi Bharucha, Nate Lee, Joe Jacobs). All of the staff across the sites who helped with logistics, planning, and operational running of the snapshot survey.

## Declaration of interests

Eight of the authors have either substantive or honorary contracts with PHE, the study funder.

**Supplement S1.** Key characteristics of SARS-CoV-2 positive HCWs.

| *SARS-CoV-2*<br>*Positive HCW* | *Age*<br>*Range* | *Gender* | *Ct value* | *Culture Status* | *Stage* | *Days since symptom*<br>*onset* |
| --- | --- | --- | --- | --- | --- | --- |
| 1 | 50-60 | F | 35.03 | Negative | Asymptomatic | - |
| 2 | 40-50 | M | 38.05 | Negative | Post-symptomatic | 22 |
| 3 | 20-30 | F | 28.18 | Negative | Post-symptomatic | 52 |
| 4 | 30-40 | F | 33.83 | Negative | Post-symptomatic | 44 |
| 5 | 40-50 | F | 32.14 | Negative | Post-symptomatic | Not recorded |
| 6 | 30-40 | M | 35.58 | Negative | Post-symptomatic | 41 |
| 7 | . | . | 26.16 | Positive | Unknown | - |
| 8 | 20-30 | F | 36.97 | Negative | Post-symptomatic | 19 |
| 9 | 20-30 | M | 29.95 | Negative | Symptomatic | 5 |
| 10 | 50-60 | M | 37.75 | Negative | Post-symptomatic | 36 |
| 11 | 50-60 | F | 39.29 | Negative | Post-symptomatic | Not recorded |
| 12 | 20-30 | M | 37.57 | Negative | Post-symptomatic | 17 |
| 13 | 30-40 | M | 36.00 | Negative | Post-symptomatic | 34 |
| 14 | 40-50 | F | 36.90 | Negative | Post-symptomatic | 39 |
| 15 | 40-50 | F | 38.30 | Negative | Post-symptomatic | 60 |
| 16 | 40-50 | F | 36.70 | Negative | Pre- or Asymptomatic | - |
| 17 | 30-40 | M | 31.22 | Negative | Post-symptomatic | 25 |
| 18 | 40-50 | F | 32.42 | Negative | Post-symptomatic | 31 |
| 19 | 60-70 | F | 36.15 | Negative | Asymptomatic | - |
| 20 | 50-60 | F | 35.68 | Negative | Post-symptomatic | 28 |
| 21 | 50-60 | M | 32.92 | Negative | Post-symptomatic | 30 |
| 22 | 30-40 | F | 37.91 | Negative | Post-symptomatic | 42 |
| 23 | 20-30 | F | 35.02 | Negative | Post-symptomatic | 13 |

## Appendix A – Questionnaire

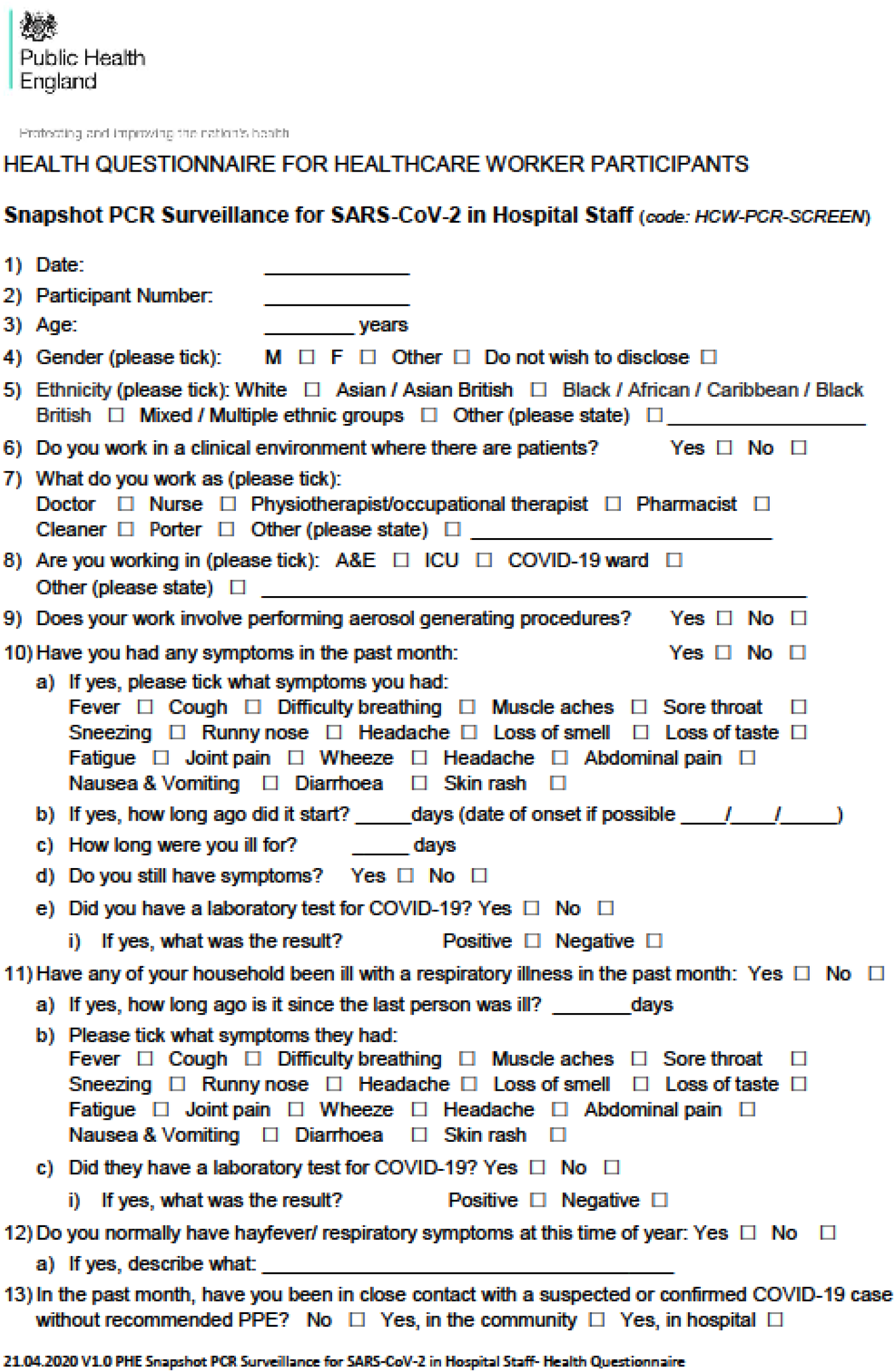

